# Enablers and barriers to scaling up an early childhood development parenting program integrated into primary health care services in rural Bangladesh: A mixed-method investigation

**DOI:** 10.64898/2026.10.28.26366714

**Authors:** Syeda Fardina Mehrin, Raafat Hassan, Masuma Kawsir, Jena Derakhshani Hamadani, Helen-Baker Henningham

**Affiliations:** Maternal and Child Health Division, icddr,b, Dhaka, Bangladesh; School of Psychology, Bangor University, United Kingdom

**Keywords:** Early childhood development, parenting intervention, primary health care services, mixed-methods, Bangladesh

## Abstract

Early childhood development programs play a crucial role in fostering child development. However, scaling-up these programs in low- and middle-income countries poses significant challenges. We conducted a mixed-method study of an evidence-based early childhood parenting program integrated into the primary health care services (community clinics) across four districts in rural Bangladesh. The program caters to caregivers of children aged 6-36 months with mild-to-moderate malnutrition. The parenting program is delivered by government community health workers (CHWs) with small groups of mother-child dyads within the community clinic setting as part of their routine duties. Mothers-child dyads attend fortnightly sessions for one year. CHWs are trained and monitored by government supervisors with overall oversight provided by government health staff at the sub-district and district level. The aim of the study was to identify the strengths and weaknesses of the program and strategies for improvement. Quantitative data included data from clinic logbooks, observations of session quality (n=60), and in-person interviews with participating mothers (n=185) in sixty clinics and phone interviews with mothers (n=164) in a further eighty clinics. Qualitative data included in-depth interviews with CHWs (n=20), government inspectors (n=16), and managers (n=4), and focus group discussions with mothers (n=4 groups, n=30 mothers), and grandmothers (n=4 groups, n=28 grandmothers) across twelve clinics. Results indicated that the key enablers to program implementation included strong government ownership, high compliance in implementing the program by CHWs, adequate session quality, satisfactory session attendance, and positive attitudes towards the program by all stakeholders. Key barriers included low levels of supervision, insufficient advocacy messaging within the community, poor communication across health staff at the different levels, and challenges in providing play materials at scale. The findings are being used to inform revisions to the program to enhance implementation fidelity and sustainability within the health system.

## Introduction

Globally, over 250 million children (43%) under the age of five in low and middle-income countries (LMICs) do not reach their full developmental potential. This is due to poverty, undernutrition, poor health, and a lack of stimulation at home [1]. These accumulated risks are associated with long-term negative effects on educational attainment and adult productivity. There is strong evidence that early childhood parenting interventions benefit young children’s development [2]. The challenge now is to scale up these interventions to extend their reach to more children at risk of poor development [1, 3]. Several studies have shown that parenting interventions integrated into existing government services can maintain effectiveness when implemented with sufficient quality and fidelity [4-7]. However, there are challenges in maintaining implementation quality including increased staff workload [8-10], staff turnover and non-compliance [8,11], low levels of supervision [10], competing priorities [10), and poor engagement of caregivers [10, 12]. There are several examples of evidence-informed interventions that have failed to improve children’s development when implemented on a larger scale [13-15]. Other large-scale studies have found only very small benefits (0.1SD) to child development [16-17]. To increase access to effective parenting services, we need a greater understanding of how these interventions can be delivered across different contexts and within different systems.

The Reach-Up parenting program, (originating from the Jamaican Early Childhood Stimulation Home Visiting Intervention), is a play-based intervention aimed at supporting mothers with children aged 0 to 48 months [18]. A meta-analysis of eighteen trials of Reach-Up across eight low- and middle-income countries found average effect sizes of 0.49 SD, 0.38 SD and 0.27 SD for child cognition, language and motor skills, respectively and an average effect size of 0.37 SD for stimulation in the home [19]. There is also evidence from a study in Jamaica that benefits are sustained over the long term, including benefits to cognition, education, mental health and earnings up to age 30 years [20-21]. In Bangladesh, we previously adapted the Reach-Up home-visiting program for integration into the primary health care services in rural Bangladesh [22]. Government health workers in community clinics delivered parenting sessions with pairs or small groups of mother-child dyads as part of their routine duties. In two cluster-randomized trials, we found positive impacts on children’s cognitive and language development, with the effect sizes ranging from 0.7 to 1.3 SD [23-24]. However, in those trials, the community health workers (CHWs) were trained and monitored by the research team. We then tested the feasibility of utilizing government health supervisors to train and supervise the CHWs to implement the parenting program, and although supervision levels were low, we found high compliance by CHWs in conducting the sessions, high levels of mother attendance and significant benefits to home stimulation with an effect size of 0.53 SD [25]. The positive findings from these studies led to the initiation of the ‘Saving Bangladeshi Babies Brains’ (SB3) program.

The aim of SB3 was to scale-up the group parenting program both vertically and horizonatally through the primary health care service. Vertical scale involved fully integrating the program into the primary healthcare service in rural Bangladesh, with responsibility for program implementation residing with the government health service. We aimed to establish an organisational structure to promote the quality and sustainability of the program when implemented on a large scale. This includes establishing training, supervision, monitoring and reporting protocols at the national, district, sub-district, and union levels. Horizontal scale involved extending the reach of the program by expanding the program to more districts. Mothers of children aged 6-24 months, with mild to moderate malnutrition, living in the catchment area of the clinics, are eligible to participate in the program. The SB3 program is currently operating in 613 community clinics of 21 sub-districts under four districts of Narsingdi (n=119), Habiganj (n=199), Brahmanbaria (n=200), and Lakshmipur (n=103). The latter three districts were selected due to the high prevalence of childhood malnutrition, while the first district was selected because we had conducted a previous study there. Over 10,000 mother/child dyads have participated in this program to date.

The aim of the current research was to: 1) investigate the extent to which the program was being implemented as intended within the community clinics, 2) explore the enablers and challenges to intervention implementation, and 3) use the data to identify strategies to strengthen the program.

## Methods

### Study design

We conducted a process evaluation of a large-scale implementation of the group parenting program integrated into the primary health care system in rural Bangladesh. The process evaluation focused primarily on the quality and fidelity of implementation [26]. We used a parallel mixed-method design in which quantitative and qualitative data were collected and analyzed concurrently [27]. The quantitative data included measures of the quality and fidelity of intervention implementation through structured observations, clinic records and mother surveys. The qualitative data included in-depth interviews with government health staff at different levels within the health service and focus group discussions with mothers and mothers-in-law to investigate the enablers and barriers affecting intervention implementation and suggestions for improvement. The combination of quantitative and qualitative data thus gave information ont the extent to which the intervention was implemented as intended and stakeholder perspectives of factors that affect the quality of intervention delivery and mothers’ engagement and participation in the program. Data was collected from December 2023 to May 2024.

### The context for the study

Primary health care services in rural Bangladesh are delivered through a network of community clinics. Clinics are staffed by three cadres of community health workers (CHWs): a Community Health Care Provider (CHCP) who works full time in the clinic, a Health Assistant (HA) and a Family Welfare Assistant (FWA), both of whom work 2-3 days a week in the clinic and 3-4 days a week in the community. CHCPs and HAs have Masters’ degrees; most FWAs have completed high school. At the union, sub-district and district levels, the work of the CHCPs and HAs comes under the Directorate of Health Services, while the work of the FWAs comes under the Directorate of Family Planning, both under the Ministry of Health and Family Welfare. Therefore, there are two sets of government inspectors responsible for clinic health workers: Health Inspectors supervise the CHCPs and HAs, while Family Planning Inspectors supervise the FWAs. Similarly, at the sub-district and district levels, there are Medical Officers and Health and Family Planning Officers to oversee their respective activities.

In the SB3 program, the parenting intervention is fully integrated into the health service with sessions conducted by the three cadres of CHWs within the community clinics and training and supervision provided by government inspectors at the union level (the smallest administrative unit in rural Bangladesh). The government inspectors were in turn trained and supervised by senior officers at the sub-district level with oversight of the program by district-level staff. In the intervention, mothers of children aged between 6-24 months are invited to attend fortnightly parenting sessions in groups of 4-5 mother-child dyads for one year. After one year, another set of mother-child dyads are invited to participate in the program. The core components of the intervention are: 1) encouraging parents to be responsive to their child’s vocalizations, actions, emotions and interests, 2) supporting parents to use sequenced and developmentally appropriate activities with their child that target all developmental domains but with a specific emphasis on cognitive ability and language skills, and 3) use of low-cost play materials and picture books [18]. In terms of intervention delivery, the core components include: 1) the use of demonstration and practice to introduce activities to parents, 2) positive reinforcement for both parents and children, 3) integrating activities into everyday routines and activities, 4) high quality training of facilitators and ongoing supportive supervision, 5) making sessions enjoyable for parents and for children, and 6) a focus on positive relationships at all levels of implementation. Full details of the intervention are given in Panel 1. The logic model depicting the theory of change for the intervention is shown in Figure 1. At the time this study was conducted, the intervention had been fully operational for two-to-three years, and health staff were implementing the intervention with a second or third cohort of mother-child dyads.

**Figure 1.**
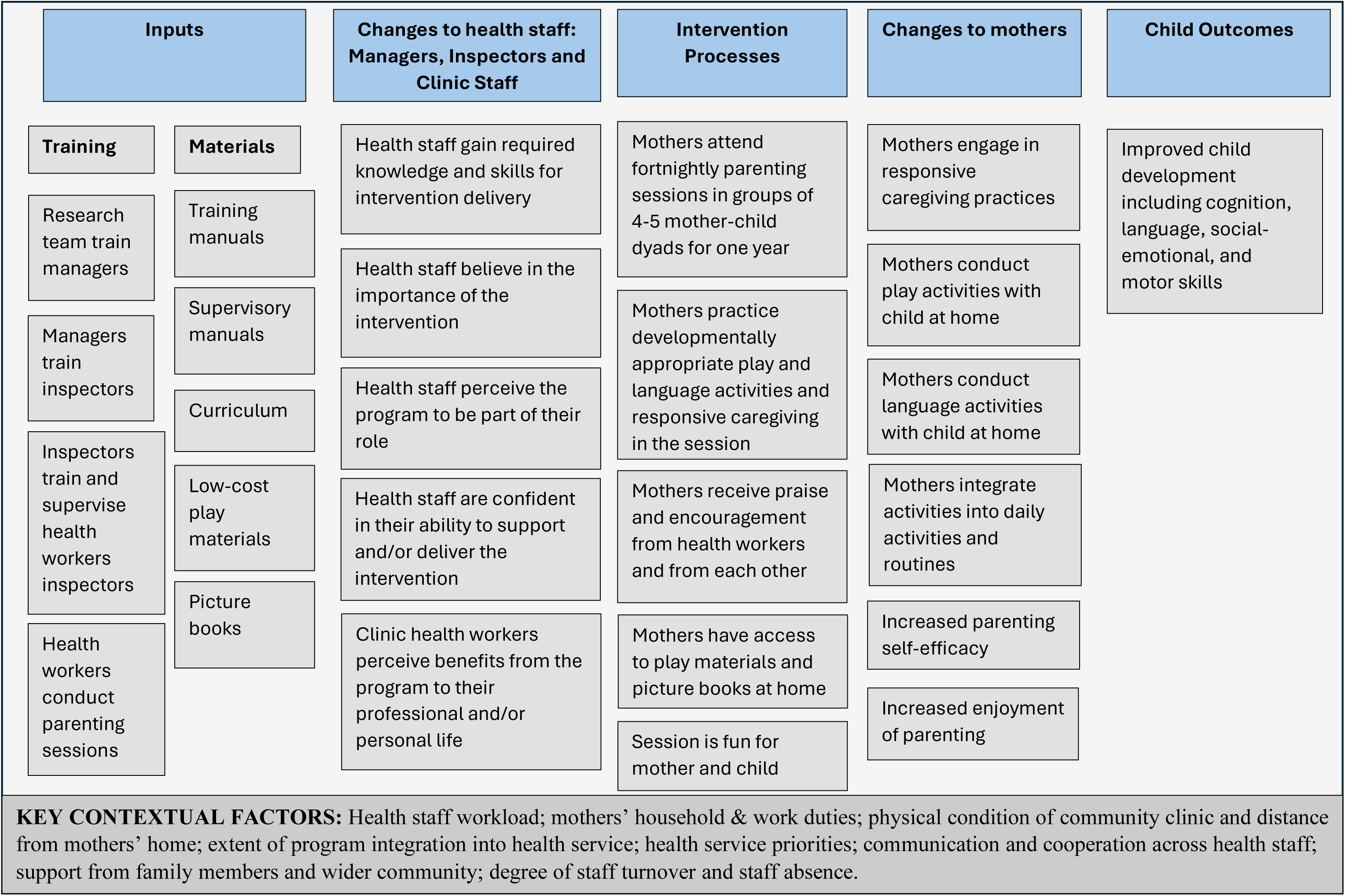
Logic Model for the Saving Bangladeshi Babies Brains (SB3) Program.

#### Panel 1. Description of Intervention.

##### Content

The curriculum is adapted from Reach-Up curriculum and focuses on promoting responsive parenting, increasing stimulation at home and increasing mothers’ parenting self-confidence. During the session, mothers are encouraged to engage in responsive, playful interactions with their child using low-cost play materials, books and materials that are available in the home, and in everyday caregiving routines. The play and book activities for children under 24 months are designed based on the concept of object permanence, verbal and physical imitation, and exploration of objects, while for children above 24 months, they are based on concepts of size, number, color, shape, position, similarities /differences and classification.

##### Materials

*Materials for Facilitators:* comprise a curriculum manual and a summary card for each session. The curriculum manual includes detailed guidelines for twelve parenting sessions which are repeated after 6 months. The summary card is a shortened version designed to be used by the Community Health Workers (CHWs) during the session. The curriculum is designed for children aged 6-36 months with activities divided into three age bands: 6-11 months, 12-23, and 24-36 months.

*Materials for Participants:* include picture books, play materials made from recycled materials (e.g. shakers, stacking toys, push-a-long toys, nesting toys), wooden blocks, soft toys (e.g. doll, ball, bean bag), puzzle boards, matching games, and crayon and paper. Mothers are given an age-appropriate toy and a book at each session, and these are exchanged with a different toy and book at the next session.

##### Procedure

Mothers attended the sessions with their child in groups of 4-5 mother-child dyads. Sessions were conducted at community clinics. Each session includes the following activities: 1) feedback from the previous session, 2) a song, 3) demonstration and practice of a toy, book and language activity, 4) a nutritional or developmental message, and 5) review and reminder of home activities. During the session, the CHWs demonstrate each activity and then support mothers as they practice the activity with their child. Mothers are also encouraged to share ideas for other activities and songs. There is a strong focus on building positive relationships with the mothers, using praise and encourage to increase their parenting self-confidence encouraging mothers to praise their children.

##### Staffing

*Facilitators:* 1,801 frontline government CHWs, including 568 Community Health Care Providers (CHCPs), 571 Family Welfare Assistants (FWAs) and 662 Health Assistants (HAs)) from 613 community clinics were trained to conduct the parenting sessions. All FWAs, 388 (56%) CHCPs, and 346 (44%) HAs were female. They received 5 days of initial training in groups of 12-15 participants and a 1-day refresher training every 6 months. An honorarium was provided for each day of training attended according to the established government rates. A fully structured training manual was used for all training.

*Trainers:* We used a cascade model of training. The research team trained medical officers and family planning officers (n=64) at the district level who then trained the government health inspectors and family planning inspectors (n=421) at the sub-district level. The inspectors then trained the frontline health workers.

*Supervision:* Government inspectors are tasked with monitoring the SB3 activities in the clinic as part of their routine duties. This involves observing a session conducted by each CHW once a month. Inspectors report to medical officers and family planning officers.

##### Duration and frequency

Each mother-child dyad was invited to participate in fortnightly parenting sessions over the period of one year. Session duration was 45-50 minutes.

### Study sample

#### Sample for observations of session quality

Within each district, one field coordinator has responsibility for monitoring program activities. Each field coordinator was asked to identify clinics within their respective district that had started intervening with a new cohort of mother-child dyads more than four months ago. From those clinics, each field coordinator provided a list of 60 clinics classified into three groups: high-performing clinics, medium-performing clinics, and low-performing clinics. Field co-ordinators based their judgements on their interactions with clinic staff including their level of cooperation, timeliness in submitting reports and engagement with the program. A total of 240 clinics were included in the listing (sixty per district). We used stratified random sampling to select a total of sixty clinics, with twenty clinics in each category. We selected twenty clinics in both the Habiganj and Brahmanbaria district and ten clinics in Narshingdi and Lakshmipur district, proportionate with the number of the clinics in each district. Within each clinic, we observed one parenting session (n=60) to evaluate the quality of implementation.

#### Sample for mother interviews at home

From each of the community clinics selected for the observational sample (n=60), we randomly selected four mothers who were currently enrolled in the program. Interviewers were requested to schedule interviews with three of the four mothers with one mother acting as a reserve. A total of 185 mothers were interviewed: 60 from Habiganj district, 60 from Brahmanbaria, 32 from Narshingdi and 33 from Lakshmipur.

#### Sample for mother interviews by phone

Phone interviews were conducted with mothers in an additional eighty clinics to extend the number of clinics included. Twenty clinics from each of the four districts were selected including forty high-performing and forty medium-performing clinics. We dropped low-performing clinics as it was difficult to collect mothers’ phone numbers. We collected the list of mothers’ phone numbers from the CHWs and phone interviews were conducted with two-to-three mothers from each clinic. If a mother was unavailable, they called the next mother in the list. A total of 164 mothers were interviewed over phone: 31 from Habiganj district, 46 from Brahmanbaria, 45 from Narshingdi and 42 from Lakshmipur.

#### Sample for qualitative interviews and focus group discussions

From the sixty clinics participating in the observations, we selected a purposive sample of twelve clinics to participate in an in-depth qualitative investigation representing high, medium and low performing clinics across all four districts.

In-depth interviews were conducted with health staff including community health workers, government inspectors and health managers. Within each of the twelve clinics, we selected one or two community health workers and within each district, we invited four inspectors to participate in in-depth interviews. A total of twenty community health workers participated: eight Community Health Care Providers, seven Health Assistants and five Family Welfare Assistants. Sixteen inspectors, including twelve Health Inspectors and four Family Planning Inspectors were interviewed. We also conducted in-depth interviews with five, district-level managers including three Family Planning Officers and two Medical Officers.

Focus group discussions were conducted with mothers and grandmothers. We invited mothers from four high-performing clinics (one per district) and mothers-in-law from four mid- and low-performing clinics (one per district) to participate in focus group discussions. We recruited mothers from high-performing community clinics to allow us to investigate perspectives of mothers who were more likely to be knowledgeable about the program. We selected mothers-in-law (grandmothers) from mid to low performing CCs to understand potential challenges with program implementation within the community. We have included grandmothers as they are the primary gatekeepers to mothers’ participation. Mothers and grandmothers were identified with the assistance of the respective CHWs. A convenience sampling approach was used based on the availability of the participants. In each FGD, there were six to eight participants. We conducted four FGDs with a total of thirty mothers and four FGDs with a total of 28 grandmothers across the four intervention districts.

### Ethical approval

Ethical approval for the study was given by the Institutional Review Board of the International Centre for Diarrhoeal Disease Research, Bangladesh (icddr,b) (Approval No IRB 19040). Written informed consent was obtained from all participating caregivers, grandmothers, CHWs, inspectors and managers.

### Measurements

In this mixed-method investigation we collected three main categories of data: 1) the quality and fidelity of intervention implementation through observation of parenting sessions and from clinic records, 2) mother reports on how sessions are conducted and enactment of the activities at home, and 3) stakeholder perspectives of the enablers and barriers to implementation and suggestions for improvement. Details of these measures are given in Panel 2.

#### Panel 2. Measurements Used in the Study.

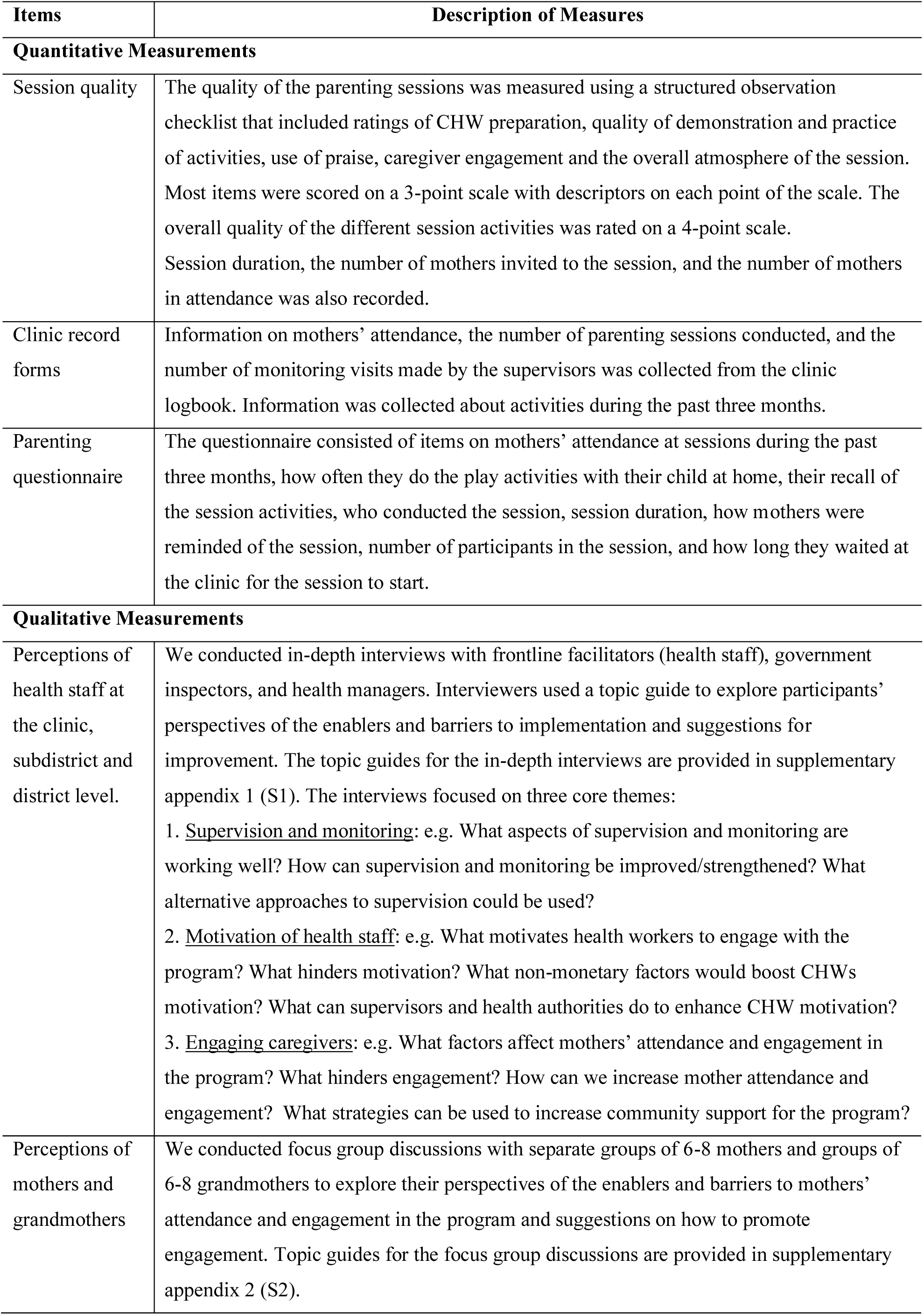

| Items | Description of Measures |
| --- | --- |
| **Quantitative Measurements** |  |
| Session quality | The quality of the parenting sessions was measured using a structured observation checklist that included ratings of CHW preparation, quality of demonstration and practice of activities, use of praise, caregiver engagement and the overall atmosphere of the session. Most items were scored on a 3-point scale with descriptors on each point of the scale. The overall quality of the different session activities was rated on a 4-point scale. Session duration, the number of mothers invited to the session, and the number of mothers in attendance was also recorded. |
| Clinic record forms | Information on mothers' attendance, the number of parenting sessions conducted, and the number of monitoring visits made by the supervisors was collected from the clinic logbook. Information was collected about activities during the past three months. |
| Parenting questionnaire | The questionnaire consisted of items on mothers' attendance at sessions during the past three months, how often they do the play activities with their child at home, their recall of the session activities, who conducted the session, session duration, how mothers were reminded of the session, number of participants in the session, and how long they waited at the clinic for the session to start. |
| **Qualitative Measurements** |  |
| Perceptions of health staff at the clinic, subdistrict and district level. | We conducted in-depth interviews with frontline facilitators (health staff), government inspectors, and health managers. Interviewers used a topic guide to explore participants' perspectives of the enablers and barriers to implementation and suggestions for improvement. The topic guides for the in-depth interviews are provided in supplementary appendix 1 (S1). The interviews focused on three core themes:<br>1. <u>Supervision and monitoring</u>: e.g. What aspects of supervision and monitoring are working well? How can supervision and monitoring be improved/strengthened? What alternative approaches to supervision could be used?<br>2. <u>Motivation of health staff</u>: e.g. What motivates health workers to engage with the program? What hinders motivation? What non-monetary factors would boost CHWs motivation? What can supervisors and health authorities do to enhance CHW motivation?<br>3. <u>Engaging caregivers</u>: e.g. What factors affect mothers' attendance and engagement in the program? What hinders engagement? How can we increase mother attendance and engagement? What strategies can be used to increase community support for the program? |
| Perceptions of mothers and grandmothers | We conducted focus group discussions with separate groups of 6-8 mothers and groups of 6-8 grandmothers to explore their perspectives of the enablers and barriers to mothers' attendance and engagement in the program and suggestions on how to promote engagement. Topic guides for the focus group discussions are provided in supplementary appendix 2 (S2). |

#### Quantitative data collection

Five research assistants (RAs) who were not involved in intervention implementation and with no prior contact with the community clinics collected the quantitative data. All had prior experience in collecting similar data within the health system in previous studies. The RAs were trained by SFM and MK who have extensive experience in collecting observational data and administering parent questionnaires in community clinic settings. RAs were trained for five days in the use of the observation checklist and clinic record form to measure the quality and fidelity of implementation. They were trained for an additional five days in the administration of the parenting questionnaires. Each training comprised three days in-office training followed by two days of field practice. Observations were conducted within the community clinic during a routinely scheduled session and data was collected using paper and pencil methods. RAs completed the structured observational checklist based on their observations of the entire parenting session and then used the clinic record book to record the data on mother attendance, number of sessions conducted and supervisory visits. Parenting questionnaires were administered using pencil and paper methods in a face-to-face interview at the participant’s home or by phone. The average duration of the interview was 20 minutes for phone interviews and 30 minutes for face-to-face interviews.

#### Qualitative data collection

The qualitative research team comprised four male and four female RAs with a Masters in anthropology and experience in conducting in-depth interviews and focus group discussions, including in health settings. The RAs had no prior connections with the participants and were new to the field of early childhood development. In-depth interviews were conducted by both male and female members of the qualitative team. The focus group discussions were facilitated by the female members of the team only. The team was trained over two weeks by the qualitative lead, with support from the intervention lead, through a combination of in-office training followed by field practice with stakeholders from areas not selected for the qualitative investigation. The interviews with the managers and inspectors were conducted at the sub-district hospital, their designated duty stations. The interviews with CHWs and the focus groups with mothers and grandmothers were conducted at their respective community clinics. The average duration of interviews with managers was 35 minutes, with inspectors 50 minutes, and with CHWs approximately 60 minutes. The focus group discussions averaged 50 minutes in duration. Interviewers made debrief notes on the main themes emerging after each in-depth interview and focus group discussion and these notes were used during weekly meetings with the qualitative lead to identify emerging themes and to promote quality control. All interviews were audiotaped and transcribed verbatim and then the transcripts were independently checked for accuracy against the audiotape.

### Data analysis

Quantitative data was entered into SPSS and analysed using descriptive statistics. Qualitative data was analysed manually using thematic analysis [29]. We used the individual topic guides to develop a list of key themes and subthemes for each type of respondent (CHW, inspector, manager, mother, grandmother) using a deductive approach. Additional codes were added inductively as new themes emerged from the data. We developed a codebook in which each code was described in detail along with examples of quotes that matched each code. To ensure intercoder reliability, the team of eight qualitative RAs engaged in a week-long training session with the lead qualitative researcher that involved discussion of the codes and hands-on coding exercises. Finally, two rounds of coding were conducted in which each of the transcripts were coded by two separate RAs. The lead qualitative researcher reviewed the coded transcripts, and any discrepancies were discussed and resolved within the team. Analyses were conducted separately for each type of respondent, and the results were combined in a final stage to identify commonalities and differences across respondents. All analyses were conducted with the transcripts in Bangla and once analyses were complete, key quotes were identified and translated to English.

## Results

### Quality of implementation of parenting session

Sixty CHWs were observed conducting a parenting session. There was a mean (range) of 7.55 (3-20) mother-child dyads in attendance and sessions lasted a mean (range) of 37.2 (14-60) minutes. Mothers waited a mean (range) of 13.7 (5-40) minutes for the sessions to start. The quality of the sessions was adequate across most indices (Table 1).

**Table 1.**
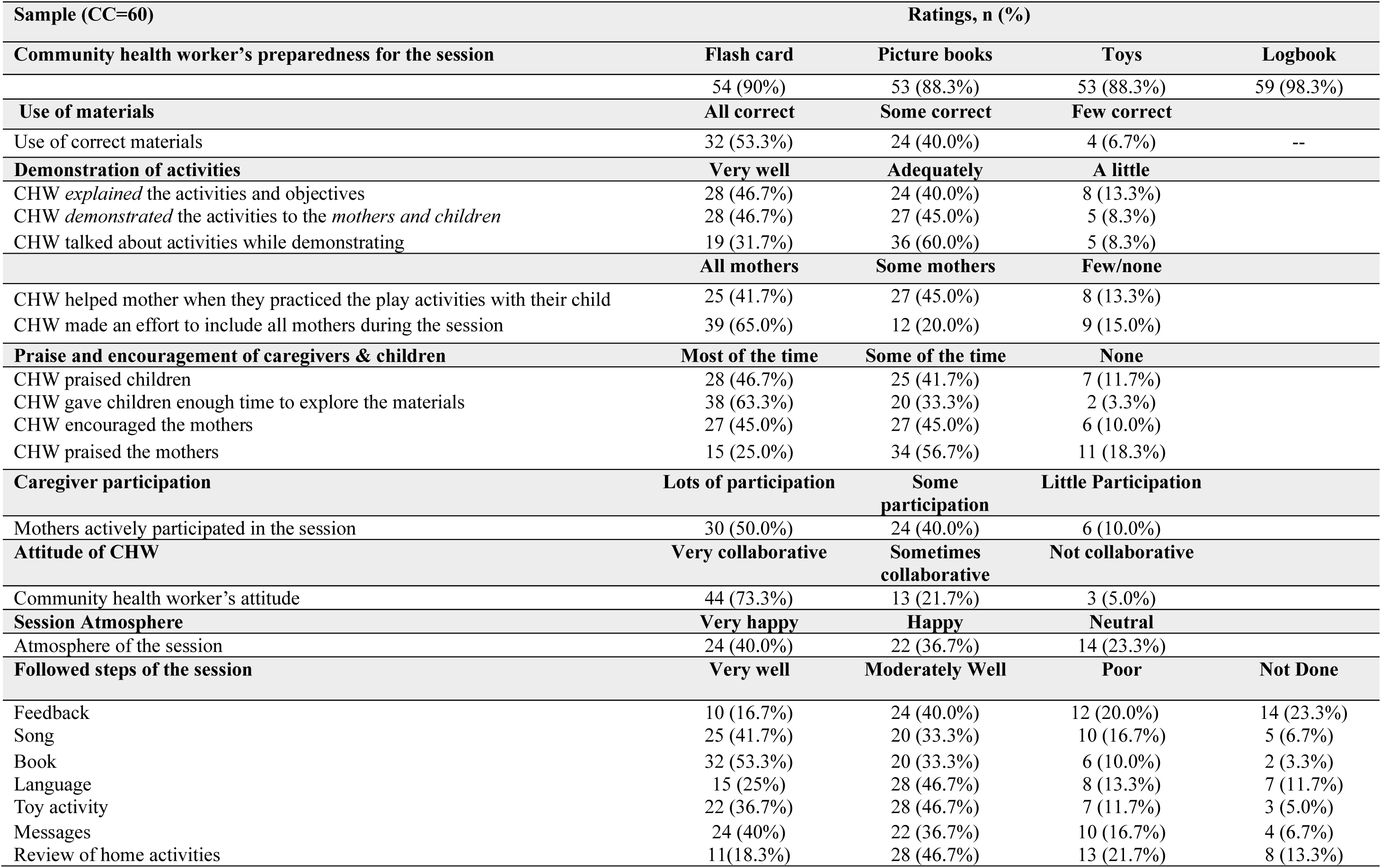
Observed Quality of Parenting Sessions.

Relative strengths (in which at least 50% of sessions were rated high) were CHW’s level of preparedness for the session, use of correct materials, including all caregivers in session activities, giving time for children to explore the materials, caregivers’ actively participating in the session, and the CHW using very collaborative methods during the delivery of the books and toy activities. The main weaknesses were related to CHW’s lack of praising the mothers and poor-quality delivery and/or omission of the feedback, language activity and review parts of the session. CHW’s compliance in conducting sessions was high with 56 out of 60 (93.3%) conducting all six sessions in the past three months (Table 2). However, supervision was low with only 11 out of 60 (18.3%) receiving the required three supervisory visits in the past three months and 32 out of 60 (53.3%) receiving no visits (Table 2).

**Table 2.** Number of Sessions Conducted and Number of Supervisory Visits.

| **Number of sessions conducted by health workers in past 3 months** | **N (%)** |
| --- | --- |
| Number of health workers who conducted 6 sessions in past 3 months | 56/60 (93.3%) |
| Number of health workers who conducted 5 sessions in past 3 months | 1/60 (1.7%) |
| Number of health workers who conducted 3 sessions in past 3 months | 2/60 (3.3%) |
| Number of health workers who conducted no sessions in past 3 months | 1/60 (1.7%) |
| **Number of supervisory visits in past 3 months** | **N (%)** |
| Number of health workers who received at least 3 monitoring visits | 11/60 (18.3%) |
| Number of health workers who received 2 monitoring visits | 5/60 (8.3%) |
| Number of health workers who received 1 monitoring visits | 12/60 (20.0%) |
| Number of health workers who received no monitoring visits | 32/60 (53.3%) |

### Results from questionnaire with mothers

#### Face-to-face interviews with mothers

Mothers reported high attendance at sessions with 98.4% attending at least five out of six sessions (Table 3). Mothers were asked to state the main reasons for attending sessions and the most common reasons were finding the sessions beneficial for children (82.1%), their child enjoying the activities (66.3%) and receiving important information on taking care of their child (51.6%). Most mothers recalled the details of the song, toy, and book activity and the developmental or nutritional messages; however, fewer mothers (38.1%) recalled the language activity. Most sessions were conducted by the CHCP, who is the core person at the clinic, although 35.3% reported that another CHW assisted during the session. Most mothers (>90%) were reminded of sessions by phone calls or home visits from the CHW; only a few followed the calendar provided by the clinic. Most mothers (95%) reported waiting less than thirty minutes for sessions to start. More than 95% of the mothers reported that they did the activities with their children at home, and more than 56% reported playing with their child every day. Mothers also reported that fathers and other family members did the activities with the child at home. The program offers a toy library in which toys and books are given to each mother-child dyad during the session and then returned and swapped for a new toy and book at the next session. However, 29% of mothers reported that they did not return any of the materials.

**Table 3.** Results from Face-to-Face Interviews with Mothers.

| **(n=185 mothers)** | **N (%)** |  |  |  |  |
| --- | --- | --- | --- | --- | --- |
|  | **5 times** | **6 times** | **Not attended** |  |  |
| # of sessions attended in the past 3 months (n=183)* | 91 (49.7%) | 91 (49.7%) | 1 (0.55%) |  |  |
|  | **Child enjoys activities** | **Session is fun** | **HW is friendly** | **Important knowledge about child care** | **Beneficial for children** |
| Stated reasons why mothers attended the sessions | 122 (66.3%) | 17 (9.2%) | 23 (12.5%) | 95 (51.6%) | 151 (82.1%) |
|  | **Song** | **Toy** | **Language** | **Book** | **Messages** |
| Recall of the activities of the session by mothers | 156 (84.3%) | 176 (95.1%) | 72 (38.1%) | 164 (88.9%) | 184 (99.4) |
|  | **< 30 minutes** | **30 to 60 minutes** | **> 1 hour** |  |  |
| Recall of the duration of the session by mothers (n=173)* | 12 (6.9%) | 102 (59.0%) | 59 (34.1%) |  |  |
| Mother's waiting time at the clinic before session start (n=128)* | 121 (94.5%) | 4 (3.1%) | 3 (2.3%) |  |  |
|  | **CHCP** | **FWA** | **HA** |  |  |
| Who conducted the sessions (n=169)* | 158 (93.5%) | 3 (1.8%) | 8 (4.7%) |  |  |
|  | **Calendar** | **Over phone** | **Home visit** |  |  |
| How mothers are reminded about the sessions (n=172)* | 16 (9.3%) | 120 (69.8%) | 36 (20.9%) |  |  |
| **Practice of activities in the past week** | **Everyday** | **3-4 days per week** | **1-2 days per week** | **No** |  |
| Mother played with child at home using the toys | 105 (56.7%) | 68 (36.8%) | 4 (2.2%) | 8 (4.3%) |  |
| Father played with child at home | 29 (15.7%) | 40 (21.6%) | 44 (23.8%) | 69 (37.3%) |  |
| Other family members played with child at home | 98 (53.0%) | 46 (24.9%) | 25 (13.5%) | 12 (6.5%) |  |
|  | **1 to 5 mothers** | **7 to 10 mothers** | **> 10 mothers** |  |  |
| Number of mothers attending last session (n=172)* | 41 (23.8%) | 115 (66.9%) | 16 (9.3%) |  |  |
|  | **All materials** | **Some materials** | **Not returned any** |  |  |
| Mother returned the toys | 117 (63.2%) | 14 (7.6%) | 54 (29.2%) |  |  |
| *Toys received: Mean (range)* | 5.1 (0-24) |  |  |  |  |
| *Books received: Mean (range)* | 5.86 (0-16) |  |  |  |  |
|  | **Yes** | **No** |  |  |  |
| Other HW helped in the session (n=173)* | 61 (35.3%) | 112 (64.7%) |  |  |  |

\*Numbers are <185 as some mothers answered don't know or don't remember

#### Phone interviews with mothers

Nearly 57% of mothers reported attending both sessions in the past month, with only 16% attending neither session (Table 4). The main reasons for not attending sessions were due to sickness of the child or mother and the mother not having time to attend. Similar to the results from the face-to-face interviews, mothers were more likely to recall the toy and book activity and the developmental or nutritional messages and least likely to recall the language activity. Mothers reported that most sessions were conducted by the CHCP, although it was common for another CHW to assist. Most mothers (>77%) were reminded of sessions by phone calls or home visits from the CHW; only a few followed the calendar provided by the clinic or were reminded through the previous meeting. Most mothers reported waiting less than thirty minutes for the session to start and most sessions were reported to be more than one hour in duration. Nearly 84% of mothers reported a group size of more than seven mother-child dyads and nearly 55% reported a group size of more than ten. Over 20% of mothers did not receive a toy in the previous session and 57% reported not returning play materials to the clinic.

**Table 4.**
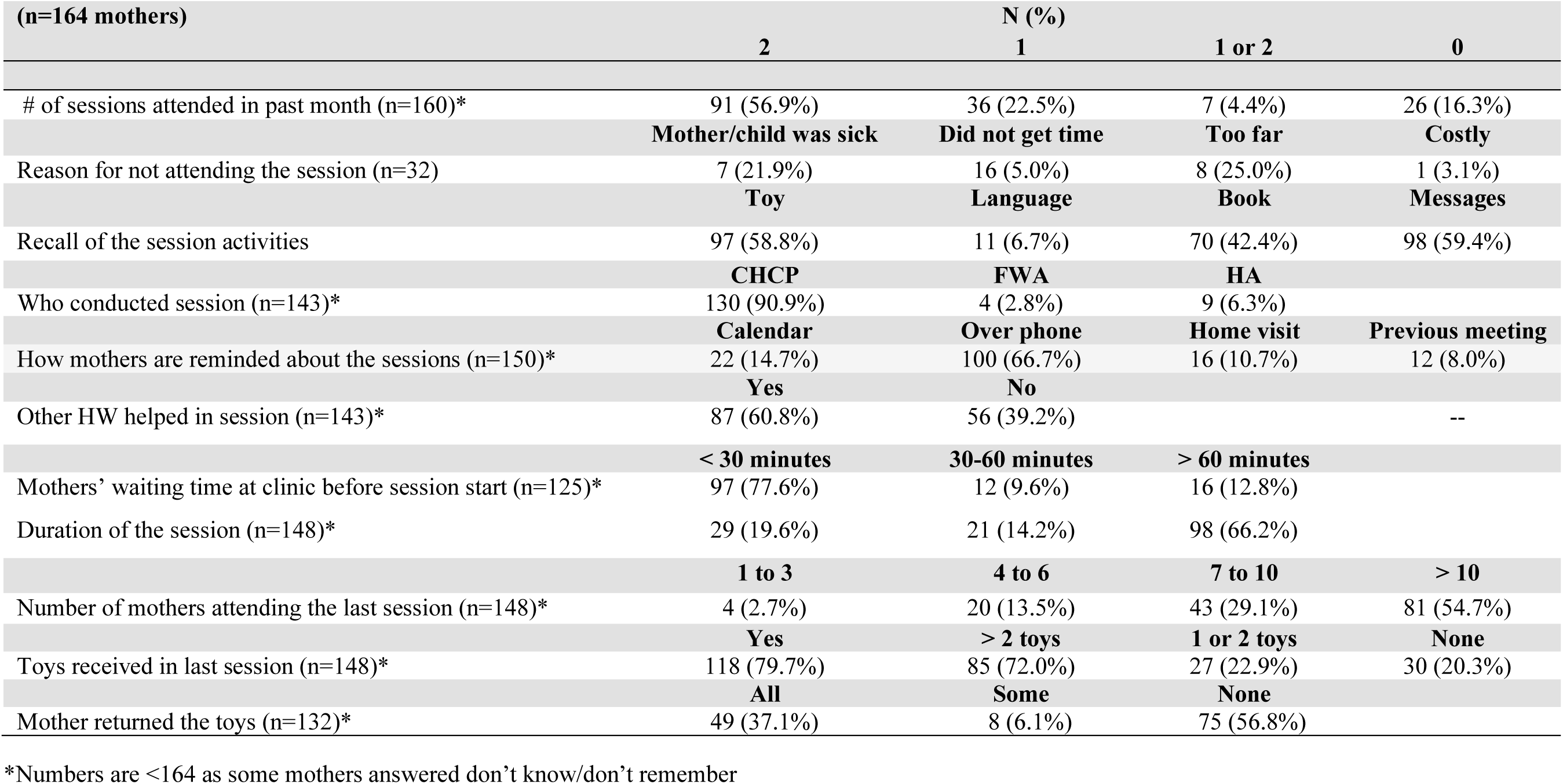
Results from Phone Interviews with Mothers.

### Qualitative results

#### Engaging caregivers in the intervention

We identified common themes across the different types of respondents relating to the enablers and barriers to caregiver attendance at sessions and engagement in the program (Table 5). An important enabler to caregivers’ participation was the observable benefits to the children from attending the sessions and these benefits were observed by mothers, grandmothers and other family members. Another enabler was the efforts by the CHWs to inform the community about the importance of the program during their routine duties.

**Table 5.**
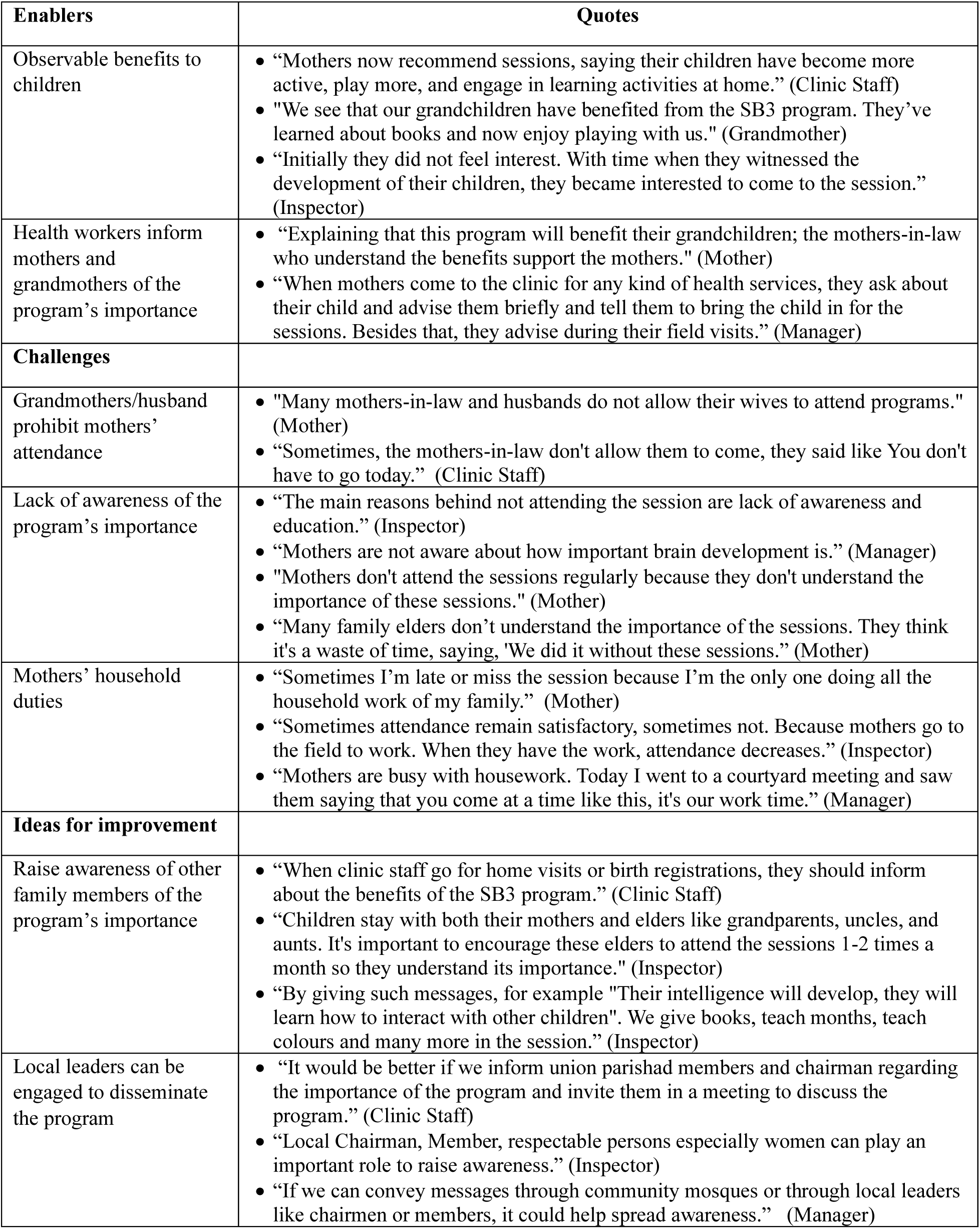
Stakeholders’ Perceptions of the Enablers, Challenges, and Suggestions for Improvement Related to Mothers’ Engagement.

Conversely, a lack of understanding of the importance of the program within the community acted as a barrier to participation and mothers were prevented from attending sessions by prohibitions from their husband and mother-in-law and because of the demands of their household duties. Participants suggested that CHWs need to continue to raise awareness of the importance of the program among all family members during routine clinic and home visits. They also suggested that influential community members could be engaged to promote the program. This would include religious leaders like imams, schoolteachers, and community leaders, in addition to health staff.

#### Motivation of community health workers (CHWs)

CHWs, inspectors and managers were asked about their perspectives on the enablers and barriers to CHWs’ motivation to engage with the program. The most commonly mentioned enablers were the CHWs receiving acknowledgement and praise for their active involvement in the program from their supervisors and the fact that the CHWs received positive feedback from mothers and observed positive changes in the participating children (Table 6). The inspectors reported that visits by district managers to observe program activities acted as a motivator for the CHWs.

**Table 6.** Stakeholders’ Perceptions of the Enablers, Challenges, and Suggestions for Improvement Related to Community Health Workers’ Motivation.

| **Enablers** | **Quotes** |
| --- | --- |
| Motivational words from higher authorities | • “We are working hard, and when they say that the session went really well and encourage us, it makes us very happy.” (Clinic Staff)<br>• “If they get a well done then they feel good. These act as motivation.” (Inspector) |
| Visits from management | • “To make them feel importance of the program, officials should visit more frequently.” (Inspector) |
| Seeing benefits to children and receiving positive feedback from mothers | • “We are doing a good deed to my community which is a religious duty.” (Clinic Staff)<br>• “I feel greatly motivated when people appreciate me as I taught them something important.” (Clinic Staff)<br>• "If the children’s brains improve playing with these toys, if we can contribute in this regard, their family will be greatly benefitted. " (Inspector)<br>• “I told them in every training session, when this program will be established nationally, the nation will remember you with gratitude.” (Manager) |
| **Challenges** |  |
| Lack of provisions for phone calls and refreshments | • “Though this is the government's responsibility, we have to cover costs like phone calls from our own pockets.” (Clinic Staff)<br>• “They (CHW) say, 'Sir, we have to spend from our pockets and conduct the sessions with our own money.’” (Manager) |
| Workload and manpower shortages | • “Our workload is increasing day by day, but the opportunities, benefits, and other necessary resources are not increasing accordingly. (Clinic Staff)<br>• “During the session we stop our regular activities. It is not possible for CHWs to do two things at the same time alone.” (Clinic Staff)<br>• “The program will be more successful if we can Increase staff. We cannot do our work properly due to lack of manpower.” (Inspector) |
| Lack of supervision | • “We are not receiving any feedback so we are missing out on important advice or guidance that could help improve our work.” (Clinic Staff) |
| **Ideas for improvement** |  |
| Additional training for community health workers | • “When we receive the training, we start our work with renewed enthusiasm and now I can identify the areas where I have flaws to improve them. (Clinic Staff)<br>• “Refreshers should be given every now and then. Then CHWs can understand the importance of the work.” (Inspector)<br>• “Refresher training can be provided to motivate health workers, as they enjoy the training and receive additional remuneration for participating.” (Manager) |
| Awards and recognition for community health workers | • “It would motivate me if my hard work was recognized. I will be happy if they give me thanks and appreciate me when I perform well.” (Clinic Staff)<br>• "If someone gave a certificate, saying, "For your good work, here's a paper." Or they gave a pen, saying, "You received a pen for your good work." It may not be anything big, but when it's given in front of everyone, it's a form of recognition. That person will feel more valued". (Inspector) |
| Regular supervision and monitoring | • “When the supervisor comes to observe the session, it motivates us.” (Clinic Staff)<br>• “Supervisors should be more willing to assist with the program.” (Inspector)<br>• “To make them feel the importance of the program, officials should visit more frequently.” (Inspector)<br>• “Observation and supervision are more important. To make the whole system successful, active involvement is necessary.” (Manager) |
| Director General mandates inclusion of program in CHW role | • "If the SB3 program is made mandatory for us by higher authority then it will become part of the regular routine for us." (Clinic Staff)<br>• “The SB3 activities should be integrated into the routine tasks. This should be made mandatory through senior officials. (Inspector)<br>• “The program needed to be included in the daily routine of the clinic staff and supervisors. I cannot do it by myself. DG should instruct us to include it in our daily routine. The instruction required to come from that level.” (Manager) |

The main barriers were resource constraints. CHWs were not provided with funds for phone calls or refreshments and some CHWs used their own personal funds to support program activities. Heavy workloads and insufficient staffing in some clinics reduced CHWs’ motivation and some CHWs reported being demotivated due to the lack of support and supervision from their supervisor.

Ideas for improvement were common across all types of health staff. A commonly mentioned theme was providing additional training to CHWs to maintain their motivation. The training is highly valued as it is enjoyable, useful, and is also remunerated according to government daily rates. Providing awards and recognition to CHWs, including verbal praise, certificates and/or small prizes were suggested. CHWs, inspectors and managers acknowledged the importance of regular, supportive supervision and reported a need to strengthen the supervisory mechanisms. Finally, participants suggested including the SB3 program as a core part of the CHWs’ role.

#### Supervision and monitoring

CHWs valued the visits from their supervisors and reported that the visits helped them to improve their skills in session delivery (Table 7). Supervisory visits were most likely to take place when the CHW and inspectors liaised together about the session schedule and/or planned the schedule together. Clinic staff reported that conducting the sessions together in a collaborative way was helpful and led to improved delivery. Inspectors and managers also reported on the importance of good communication among the various cadres of health staff, including integrating updates on the SB3 activities in the monthly meetings with CHWs, and inspectors sending regular reports on program activities to their manager so they can follow-up as necessary.

**Table 7.** Stakeholders’ Perceptions of the Enablers, Challenges, and Suggestions for Improvement Related to Monitoring and Supervision.

| **Enablers** | **Quotes** |
| --- | --- |
| Supervisors' presence and support | • "Supervisors tell us their experiences following the session. Hearing this helps me to get better in my job. Should there be a mistake the supervisor notes, we address it in the next session." (Clinic Staff) |
| Scheduling sessions based on supervisor availability | • "CHCP tells me the session date two or three days ahead." (Inspector)<br>• "Supervisors can attend only two days in a month. We schedule sessions to ensure they can attend." (Inspector) |
| CHWs conducting sessions collaboratively | • "Sessions are more effective when we conduct the session together." (Clinic Staff)<br>• "When we conduct the session together, if any step is missed or goes off track, the other can quickly step in and add that missing part. Everything is covered and the session proceeds smoothly." (Clinic Staff) |
| Manager following up with supervisors and health workers during monthly meetings | • "Higher authorities take updates on the SB3 program during monthly meetings. They usually ask if the sessions are being conducted properly and whether the supervisors are fulfilling their duties correctly." (Inspector)<br>• "The staff send me weekly or monthly reports, and I call inspectors weekly to check how activities are progressing under their areas." (Health Manager)<br>• "I follow up with inspectors weekly and conduct follow-ups with Health Assistants and CHCPs monthly." (Manager) |
| **Challenges** |  |
| Workload and manpower shortages | • "The family planning officials, due to their administrative and field-level work visit our clinics less frequently. As a result, the monitoring of SB3 program from our family planning side is somewhat reduced." (Manger)<br>• "There is only one officer in whole district to monitor the program. Instead, one officer should be appointed in every upazila." (Clinic Staff)<br>• "Managing this program, on top of my other work, becomes an overload for me." (Manager) |
| Poor communication between staff | • "But sometimes we don't get the invitation from the CHWs. They conduct session without informing me of the session time." (Inspector) |
| **Ideas for improvement** |  |
| Additional training focused on supervision | • "If supervisor receive specialized training on supervision, the supervision system will improve." (Manager)<br>• "We could carry out the supervision and monitoring work better if we were provided more training on it." (Inspector) |
| Include supervisory visit on monthly workplan | • "If the inspectors were a bit more active and plan their visits, the work would have been done better." (Manager) |
| Engaging both health and family planning sector staff | • "If there is coordination between health and family planning sector the quality of supervision will improve." (Inspector)<br>• "The major part of SB3 falls under the Directorate General of Health Services, including CHCP and Health Assistants. FWAs are the Family Planning Services. If the authorities coordinate and work together, supervision activities can be more effective." (Manager) |
| Virtual supervision | • "Supervisors make phone calls to know about the session and also ask for session pictures to see If CHW words match the pictures." (Clinic Staff)<br>• "I can monitor the session in video call. Or if I make audio call, I can ask the CHCP update of the session and regarding mothers' attendance." (Inspector) |
| Include SB3 program in monthly meetings | • "Every month, when the report is prepared for the monthly meeting, SB 3 issues should be included. The manager will be able give suggestions." (Manager)<br>• "If higher officials want to know about the update of our work and give us some directions in weekly and monthly meeting, our supervision activity will be strengthened." (Inspector) |

The main barrier to supervision and monitoring was reported to be the workload of the inspectors and the shortage of inspectors in some subdistricts due to vacant posts. The Family Planning Inspectors were reported to be less active in supervision than the Health Inspectors. Inspectors reported that poor communication about the session schedule from some CHWs was an additional barrier.

Managers and inspectors suggested that additional training needs to be provided specifically focused on how to supervise and support the CHWs. Improved planning, better coordination between the Family Planning and Health sector staff and including the SB3 program as a standard and mandatory component of monthly reporting and monthly meetings were additional suggestions for improvement. Finally, given the difficulties faced in providing face-to-face supervision, some clinic staff and inspectors suggested using virtual supervision, by audio or video call to support the CHWs. However, these suggestions mostly related to monitoring program compliance rather than program quality.

## Discussion

In this mixed-method process evaluation, we identified both strengths and weaknesses of the implementation of the SB3 program through the primary health care services in rural Bangladesh. Importantly, CHWs showed high compliance in conducting parenting sessions, actively promoted mothers’ attendance at sessions through phone calls and during routine home visits, and were competent in using many of the required skills for session delivery.

Attendance as reported by mothers was good, and mothers were able to recall the majority of what they had learnt and reported playing with their child at home. The CHWs were less skilled in conducting activities aimed at promoting children’s language and most mothers could not recall these activities. Other concerns were that the parenting sessions were often conducted with much larger groups than recommended, play materials were not always provided for mothers to take home, and there was limited supervision by the government supervisors. From qualitative interviews with a range of stakeholders, we investigated enablers and barriers relating to caregiver engagement, health worker motivation, and monitoring and supervision. The main enabling factors affecting caregiver engagement were the mothers’ and the wider family’s belief that the program was important for promoting children’s development. Participants suggested that raising this awareness required additional efforts from both the CHWs themselves and from other influential persons within the community. In terms of CHW motivation, important influences included CHWs receiving appreciation for their work from their supervisors and from the community and receiving ongoing training and supervision. For monitoring and supervision, an important enabler was good communication and co-ordination at all levels of the health service. Many CHWs reported conducting sessions collaboratively which allowed them to support each other in implementation. Implementation barriers included insufficient staffing, insufficient resources, and staff workload which impacted many aspects of program implementation. Participants also reported that to ensure that the program is sustained, it needs to be further embedded into the health service as a mandatory component.

The positive findings of high compliance by CHWs, their efforts to promote mother attendance, and their competence in conducting sessions, alongside mothers’ recall of most of the activities and their interest and engagement in the sessions, provide evidence for the acceptability and feasibility of the program by the frontline facilitators and beneficiaries.

Sessions were mostly conducted by the Community Clinic Health Provider (CHCP) who works full-time in the clinic and who have high level of education (Masters’ degree), although other CHWs assisted at times. It is evident that the program has been adopted by the primary health care service by health staff at all levels. This relative success in integrating the SB3 program into an existing government service builds on best practices in designing, implementing and scaling interventions. For example, the SB3 program uses an evidence-based parenting program that has been carefully adapted through multiple interations for the context in which it operates [22-25]. During the adaptation process, we identified the optimal caseload for CHWs, the optimal group size, and determined the frequency and duration of the sessions taking into consideration the perspectives of both CHWs and participating mothers. The program uses a structured curriculum and structured, high-quality training focusing on demonstration and practice, features that have been identified as key to program success in prior studies [30-32]. The frontline facilitators are government CHWs, with stable employment, and who are respected and trusted by the local communities. Furthermore, the primary health care service has an existing organisational structure that can be leveraged to implement the program. Low staff turnover, respect from the community and an existing organizational structure promote the ongoing sustainment of interventions integrated into government systems [33].

The program also faces some implementation challenges. One key weakness is the low level of supervision by government inspectors. Supportive supervision with constructive feedback has been shown to increase the implementation quality of ECD parenting programs over time [34-35]. High implementation quality is in turn associated with increased participant engagement and greater benefits to parenting practices and child development [36]. In previous trials in Bangladesh, when high quality supervision was provided by the research team, large benefits to parenting outcomes and to child development and behaviour were found [23-24]. In this study, the low levels of supervision were reflected in some implementation weaknesses. Challenges in ensuring adequate monitoring and supervision has been reported in other studies and is especially difficult to achieve as interventions are scaled up [10, 32, 37]. In qualitative interviews, CHWs, supervisors and managers reported that ongoing supervision is important, not only for maintaining implementation quality, but also for promoting CHWs’ motivation and engagement with the program. The importance of CHWs feeling appreciated and valued, and gaining respect for their work, has also been shown to be key to promoting facilitator compliance and motivation in other studies [31].

Another challenge was the inconsistency in loaning the play materials to mothers for home practice and the fact that mothers did not consistently return the toys. Provision of materials requires additional manpower, funding, timely distribution, storage facilities and regular stock-taking to ensure an adequate supply of materials. These challenges in making, distributing and storing materials have been reported in other studies [30-31] and pose a significant logistical challenge to further scale-up and to sustainment within the government system.

The qualitative data also showed the importance of the demand side of the program. While the supply side incorporates what is offered by the program (such as the curriculum, process of delivery, staffing), the demand side focusses on participant enrolment, engagement and retention in the intervention [38]. In our qualitative data, respondents reported that mothers’ engagement was influenced not only by their own beliefs about the importance of the program, but also by the beliefs of other family members as mothers require permission from their husband and mother-in-law to attend the sessions. In previous trials of the program, we incorporated community awareness-raising activities to increase the demand for the program with the wider community, including the families of the beneficiaries [23-25].

These activities were omitted when moving towards scale due to their cost and complexity. Lack of advocacy on the importance of ECD at the community level is a common weakness of large-scale ECD parenting programs and it is important to identify low-cost, alternative strategies to create demand and sustain engagement for these programs [39]. Fortunately, when mother-child dyads attend the sessions, the mothers themselves and other family members observe the benefits of the program which promotes further engagement as seen in this study and elsewhere [39].

The qualitative data also highlighted the need for further strengthening of the organisational systems through which the intervention was delivered. Although there is strong political will and government buy-in for SB3 program implementation, we identified gaps in communication and co-ordination between the different levels of health staff (at community, union, sub-district and district levels) and across staff from within the Directorate of Health Services and staff within the Directorate of Family Planning. It was evident that full integration had not been achieved as the SB3 program was not routinely included in meetings and in reporting mechanisms. Full integration requires that the program becomes a core part of the role of the health service.

In table 8, we show how we have used the findings from this mixed-method study to inform revisions to the intervention. This includes addressing the challenges relating to: 1) play materials and books, 2) health staff motivation, 3) caregiver engagement, 4) program monitoring and supervision, and 5) integration of the program into the government service. In terms of the play materials and books, to address the logistical challenges of providing these materials at scale, we now include toy making demonstrations in the parenting sessions and encourage mothers to make their own toys. In addition, the picture books and blocks are for sale at low cost in village shops nearby the community clinic. For health worker motivation, we have added motivational messages and strategies to the booster training sessions for both clinic health staff and their supervisors. To promote caregiver engagement, we are utilizing key community members such as imams and schoolteachers to raise awareness about the importance of early childhood development within the community. Health staff are also encouraged to explicitly point out the benefits of the program during each parenting session. Finally, we are advocating for full integration of the program into the government health service including integrating the activities into the established reporting mechanisms, including the program as an agenda item in the scheduled monthly meetings, and including the program in the job descriptions of the health workers and inspectors. By addressing challenges across these different areas, we address the demand and supply sides of the program. The revisions to implementation aim to facilitate vertical scaling of the program within the health service and to increase the feasibility of sustainable horizontal scaling across districts.

**Table 8.** Using the Findings from the Mixed-Method Formative Assessment to Inform Revisions to the SB3 Program.

|  | **Findings** | **Suggested Actions** |
| --- | --- | --- |
| Play materials and books | • Mothers did not consistently receive the play materials required for home practice.<br>• Play materials that were loaned to mothers were not consistently returned to the clinic.<br>• Provision of the play materials and books has logistical and cost implications for the government service. | • Include demonstrations of how to make the play materials during the sessions and encourage mothers to make the toys at home.<br>• Provide one set of toys to each clinic for use as a template.<br>• Include guidance on how to teach toy making during each session.<br>• Books and blocks will be sold at low cost in shops close to the clinic. |
| Health staff motivation and engagement | • Compliance with conducting sessions was high<br>• Health staff delivered sessions with reasonable competence.<br>• Health staff are more motivated when they receive positive feedback from supervisors, observe benefits to children and families, and receive respect from the community. | • Include activities specifically targeting health worker motivation in training sessions. This includes:<br>- Explicitly pointing out the benefits of the program to mothers, children and to their own professional and personal development.<br>- Showing videos of testimonials from other health staff.<br>- Encourage health staff to reflect on the benefits of the program, how it benefits the community, and how it fits within their role.<br>• Emphasise the importance of supportive feedback with supervisors/managers. |
| Monitoring and supervision | • Supervision levels by government inspectors were low.<br>• Some activities within the parenting session were delivered with low quality or were omitted (e.g. language activities).<br>• Supervision visits are highly valued by health staff.<br>• Government inspectors need more training in how to provide supportive supervision.<br>• Effective supervision requires better co-ordination across the different levels of the health service.<br>• Health staff support each other to deliver sessions. | • Strengthen the training on the aspects of the session that were implemented with low quality for health staff and supervisors.<br>• Provide additional training for government inspectors in how to supervise health staff in program implementation.<br>• Build on the peer support modality that health staff are currently using by providing structured guidelines on how they can support each other. |
| Caregiver engagement | • Attendance and engagement of mothers requires that the immediate family (e.g. mother-in-law, father) understand the value of the program.<br>• Observable benefits to children and mothers lead to higher caregiver engagement and increased support from family members. | • Design promotional materials for use by imams, teachers and health staff.<br>• Train these community members in how to use the materials during their regular activities.<br>• Explicitly point out the benefits of the program to mothers and children during the group parenting sessions and ask mothers to identify benefits, to themselves and to their child, as a core part of each session. |
| Integration into government service | • There were reports of poor co-ordination between staff at different levels of the health service.<br>• The program is not fully integrated into the existing reporting mechanisms and routine meetings.<br>• The program has not yet been included as a mandatory part of the role of the health staff. | • Integrate the SB3 program into the community health workers' and inspectors' monthly reports on their activities.<br>• Include a discussion on the SB3 program in the monthly meetings held at the different levels of the health service.<br>• Advocate for the inclusion of the activities of the SB3 program as a core part of the role of health staff at different levels. |

The strengths of this research include the use of a mixed-method design that included quantitative and qualitative data. We used a stratified random sampling method to select the clinics for the observation and in-person interviews to reduce selection bias. In the qualitative evaluation, we gathered data from multiple stakeholders including the community (mothers and grandmothers), frontline health workers (community health workers), their supervisors, and health managers. This ensures that we are incorporating viewpoints from diverse perspectives. Furthermore, researchers collecting both quantitative and qualitative data had no previous contact with the participants and received high quality training in how to administer the measures.

There are also several limitations to the study. These limitations include the small number of observations of session quality; due to budget constraints, we were only able to observe one parenting session in sixty clinics. Furthermore, we only included 240 out of the 613 clinics in the initial sampling pool when randomly selecting clinics. This was because with only one field co-ordinator per district, there was insufficient time and resources to rate all clinics. This may reduce the representativeness of the sample. The CHWs were aware the observer would be attending, and it is possible that they were better prepared and delivered the session with higher quality when the observer was present. Social desirability bias may have influenced mothers’ responses to the interviews. In addition, some interviews were conducted by phone, and it is possible that their responses were affected by the presence of other household members. However, no sensitive questions were asked. We collected each set of data at one time point only and hence we cannot track implementation over time, and we did not have resources to get feedback from stakeholders on the interpretation of the qualitative results. The qualitative team were not experienced in early childhood parenting interventions, and this may have affected their ability to probe effectively at times. However, they received rigorous initial training and ongoing support from the qualitative lead (RH) with support from SFM (a child development expert and the intervention lead). We were also unable to include fathers in our sample and so we have no information on fathers’ perspectives of the program.

In this study, we used a mixed-method process evaluation of an ongoing, large-scale ECD parenting program, implemented through primary health care services in four districts of Bangladesh. The evaluation identified strengths and weaknesses of the program which are being used to strengthen program implementation. This includes making changes necessary to expand the reach of the program through both the supply and demand side, alongside revisions to further embed the program into the health services structures and processes. The study demonstrates the value of embedding monitoring, evaluation and learning activities into ongoing program implementation throughout the process of launching, scaling and sustaining an intervention.

## Supporting information

In-depth Interview Guidelines

Focus Group Discussion Guidelines

## Acknowledgements

We extend our sincere gratitude to all study participants, with special appreciation for the contributions of the clinic health staff. We thank all the trainers and the data collectors. The Thrive programme is funded by the UK’s Foreign, Commonwealth & Development Office (FCDO) and New Zealand’s Ministry of Foreign Affairs and Trade (MFAT), through Oxford Policy Management in collaboration with the Institute for Fiscal Studies. Thrive is a multi-country research initiative aimed at translating evidence on early childhood development into scalable, cost-effective programmes with long-term societal impact. This mixed-method study was conducted at icddr,b. We appreciate the Government of Bangladesh for granting permission to conduct the research within its health infrastructure. We also acknowledge the support and cooperation of the Directorate General of Health Services (DGHS) and the Directorate General of Family Planning (DGFP) under the Ministry of Health and Family Welfare, along with all government health staff. icddr,b is grateful to the Governments of Bangladesh and Canada for their core/unrestricted support.

## Data Availability

The quantitative datasets generated and analyzed during the current study are available upon reasonable request to the authors. The qualitative data (interview transcripts and recordings) are not publicly available due to ethical and privacy restrictions, as they contain sensitive personal information that could compromise participant confidentiality. De-identified excerpts or summarized findings may be made available upon reasonable request to the authors.

