## Supplementary material for "Enablers and barriers to scaling up an early childhood development parenting program integrated into primary health care services in rural Bangladesh: A mixed-method investigation": In-depth Interview Guidelines

### Supplementary Appendix 1
### In-depth Interview Guidelines

**Target Participants – Service Providers (Community Health Care Providers, Health Assistants, Family Welfare Assistants)**

**Key area: Supervision and Monitoring Strategies:**
**Core Aim:** In this section we will be exploring strategies to strengthen supervision and monitoring mechanisms to enhance session quality considering factors beyond financial support.
**Guided Questions:**

1. You received training and have been conducting the sessions for a while now. As you know a good session lasts about 45 minutes and during the session, you demonstrate the book, toy and language activities to the mothers using activities relevant to each child's age. And all mothers get the opportunity to practice the activities with their child while you give feedback and support.
   - To what extent are you able to conduct the sessions as you were trained?
   - If the HW reports they are doing the session well, ask, how can you tell?
   - Do you think is there any room for improvement?
   - If so, what can be done to improve the quality of the sessions? In which area particularly?
   - If the HW reports that they are not doing the session as intended, ask, why do you think you are not being able to conduct the sessions like you are supposed to? Is there any kind of support that you need? If so, what kind of support?
2. We are curious about different strategies used to organize these sessions for parents and how health workers adapt their approach to program delivery.
   - Please share how you manage scheduling, timing, and group dynamics?
   - What are the reasons for choosing one approach over another?
   - What are the challenges or benefits associated with different organizational methods?
3. We understand that everybody need some kind of support to conduct this type of sessions. Currently the inspectors have the responsibility of providing you with that support.
   - Do you think we can improve the current system that ensures that you receive the support you need?

4. Who else do you think could support you, or what other ways could be used to support health workers effectively? (Probe: probe for what kind of support they are getting if they say the support is enough)

- if they say that they are getting the enough support:
  - o Great that its working for you. We are still interested in hearing your ideas for alternative methods of supervision
- If they suggest supporting each other:
  - o How would you support each other, what would it look like, how would it work?
  - o Is there anyone else in the community, who you think can provide support?

- If they say about someone in the community,
    - Who would that be, what would they do, what would they provide?

**Key area: Increasing Mothers' Attendance:**

**Core Aim:**

- In this section we will be exploring strategies for fostering community awareness of the importance of the program to increase attendance and minimize dropout rates.

**Guided Questions:**

1. Some mothers are not attending regularly, some mothers are not coming at all, some are dropping out after a few sessions. How can we promote mothers' attendance at sessions?
    - What can you do to encourage the mothers to come consistently?
    - What can members of the community do?
    - What can the other staff in the health service do?
2. How can we better communicate the program's value within the wider community. for everyone?
    - What can we do differently?
    - How can we engage community members in understanding and supporting the program?
    - What information would resonate with both mothers and the wider community to raise awareness and encourage engagement?
3. What type of support would you need from your supervisor so that you can effectively communicate to the mothers about the value of the program?

**Key area: Motivation:**

**Core Aim:** In this section we will be exploring methods for motivating healthcare workers to deliver high-quality sessions.

**Guided Questions:**

1. With this type of work, people need to be highly motivated. Beyond financial compensation, what would make you more motivated to do this type of work?
2. What changes to the program would make you feel more invested in its success?
3. If you were tasked with promoting the program to other health workers, what aspects would you emphasize to generate excitement and prioritize participation?
4. What support from the higher authority could bolster your motivation (beyond monetary benefits)?
5. **Closing:**

Thank you for your time and dedication. Your contributions to the program's success are greatly valued. As we near the end of our interview, please feel free to share any additional insights on monitoring, and supervision strategies, raising community awareness, ensuring mother's attendance, and your motivation for the program's successful implementation.

**Target Participants – Inspectors/Supervisor (Health Inspectors, Assistant Health Inspectors, Family Planning Inspectors)**

**Key Area: Supervision and Monitoring Strategies:**

**Core Aim:** In this section we will be exploring strategies to strengthen supervision and monitoring mechanisms to enhance session quality considering factors beyond financial support.

**Guided Questions:**

1. The service providers have received the training and have been conducting the sessions for a while now. As you know a good session lasts about 45 minutes and during the session, they demonstrate the book, toy and language activities to the mothers using activities relevant to each child's age., And all mothers get the opportunity to practice the activities with their child while they give feedback and support. To what extent are the clinic staff able to conduct the sessions as they were trained?

- If the participant states that they are doing well, ask, how can you tell?
- Do you think is there any room for improvement?
- If so, what can be done to improve the quality of the sessions? In which area particularly?
- If the respondent states that the HWs are not doing the sessions as intended, ask: why do you think they are not being able to conduct the sessions like they are supposed to? Is there any kind of support that they might need? If so, what kind of support?

2. We are curious about different strategies used to organize these sessions for parents and how health workers adapt their approach to program delivery.

- How do the health workers manage scheduling, timing, and group dynamics?
- Are there any challenges or benefits associated with different organization methods?

3. Currently you have the responsibility of the supervision and monitoring of this program, and we know that you sometimes face difficulties in providing this support

- What can be done to improve the current system?

4. Do you think, you need any support from your supervisor or higher authority to conduct the supervision and monitoring more effectively? If so, what kind of support?

5. Beyond current practices and traditional inspection methods, we're interested in exploring broader approaches to monitoring and supervision that could enhance support for clinic staff. Can you share your thoughts on alternative approaches to monitoring and supervision, beyond traditional inspections that could be effective and well-received by clinic staff? What considerations would be crucial for ensuring their successful implementation?

**Key Area: Increasing Mothers' Attendance:**

**Core Aim:** In this section we will be exploring strategies for fostering community awareness of the importance of the program to increase attendance and minimize dropout rates.

**Guided Questions:**

1. Some mothers are not attending regularly, some mothers are not coming at all, some are dropping out after a few sessions.

- What do you think the Community Health Workers can do to encourage the mothers to come consistently?
- What can the community do?
- What do you think you can do?

2.. How can we better communicate the program's value within the wider community. for everyone?

- What can be done differently?
- How can community members be engaged so that they understand and support the program?
- What information would resonate with both mothers and the wider community to raise awareness and encourage engagement?

3. What type of support could you provide as a supervisor so that the health workers can effectively communicate to the mothers about the value of the program?

**Motivation:**

**Core Aim:** In this section, we will be exploring methods for motivating healthcare workers to deliver high-quality sessions.

**Guided Questions:**

1. We know that this type of work, people need to be highly motivated What steps can you take to encourage the health workers? How do you think you can be more motivated to do this type of work?
2. What changes to the program would make you and the health workers feel more invested in its success?
3. If you were tasked with promoting the program to other health workers, what aspects would you emphasize to generate excitement and prioritize participation?
4. What support from the higher authority might bolster health workers' motivation (beyond monetary benefits)?

**Closing:**

Thank you for your time and dedication. Your contributions to the program's success are greatly valued. As we near the end of our interview, please share any additional insights on monitoring, and supervision strategies, raising community awareness, ensuring mother's attendance, and your motivation for the program's successful implementation.

**Target Participants –Sub-district and District Managers (Medical Officer, UH&FPO)**

**Key area: Supervision and Monitoring Strategies:**

**Core Aim:** In this section we will be exploring strategies to strengthen supervision and monitoring mechanisms to enhance session quality considering factors beyond financial support.

**Guided Questions:**

1. We recognize the importance of regular supervision and monitoring for the program's success. Please share your thoughts on how we can improve the supervision and monitoring system?
2. What role can you play to strengthen the current supervision system? Do you think, you need any support for successful implementation of the program? If so, what kind of support?

**Key area: Increasing Mothers' Attendance:**

**Core Aim:** In this section we will be exploring strategies for fostering community awareness of the importance of the program to increase attendance and minimize dropout rates.

**Guided Questions:**

1. Some mothers are not attending regularly, some mothers are not coming at all, some are dropping out after a few sessions. What do you think are the biggest barriers preventing mothers from participating consistently? Is there anything the Community Health Workers, Inspectors or members of the community itself could do to make the program more accessible and engaging for them? What do you think you yourself as a manager can do?
2. How can we better communicate the program's value within the wider community. for everyone?
   - How can we engage community members in understanding and supporting the program?
   - What information would resonate with both mothers and the wider community to raise awareness and encourage engagement?

**Motivation:**

**Core Aim:** In this section we will be exploring methods for motivating healthcare workers to deliver high-quality sessions.

1. We know that this type of work, people need to be highly motivated, beyond monetary benefits, how do you think the health workers and inspectors can be more motivated to do this type of work? What steps can you take to encourage them?
2. What changes to the program would make the health staff feel more invested in its success?
3. If you were tasked with promoting the program to health workers/inspectors, what aspects would you emphasize to generate excitement and prioritize participation?

**Closing:**

Thank you for your time and dedication. Your contributions to the program's success are greatly valued. Please share any additional insights on monitoring, and supervision strategies, raising community awareness, promotng mother's attendance, and increasing staff motivation.
