## Supplementary material for "Enablers and barriers to scaling up an early childhood development parenting program integrated into primary health care services in rural Bangladesh: A mixed-method investigation": Focus Group Discussion Guidelines

### Supplementary Appendix 2
### Focus Group Discussion Guidelines

**Target Participants: Caregivers, Mothers**

**Key Area: Increasing Mother's Attendance**
**Core Aims:**

- To develop strategies for fostering community awareness of the importance and value of the program's significance to mothers, motivating consistent attendance and minimize dropout rates.
- To investigate how health workers can effectively communicate the value of the intervention (including the provided books and toys) to the mothers, the families and the wider community.

**Guided Questions:**
We're trying to find out how to encourage mothers to engage with the program consistently and understand how we can better communicate the program's value to you and other mothers.

1. One issue we face with the program is that some mothers are not attending regularly, some mothers are not coming at all, and some are dropping out after a few sessions,
    - What are your thoughts on why mothers might not be attending the sessions regularly?
    - Can you share any personal experiences or observations that might help us understand this better?
2. What could the health workers do to encourage mothers to participate in the sessions?
3. We understand that some mothers prefer the toys they buy from the market for their child. Although these toys used in this program are very simple and homemade, they have been specifically designed to help your child to learn and do better in school. How can we convey the message that these toys are helpful/beneficial?
4. We are aware of the limited awareness about our program's importance within the wider community. How can we better communicate the program's value for everyone?
    - What can we do to raise community awareness?
    - How can we engage community members in understanding and supporting the program?
    - What information would resonate with both mothers and the wider community to raise awareness and encourage engagement?
5. Let's imagine we are designing a campaign to increase attendance and awareness. How can we make the program more appealing and accessible to the community?

**Closing:**
Thank you all for your time. As we near the end of our discussion, please feel free to share any additional insights on raising community awareness and ensuring mother's attendance for the program's successful implementation.

**Target Participants: Mothers-in-Law**

**Key Area: Increasing Mother's Attendance**
**Core Aims:**

- To develop strategies for fostering community awareness of the importance and value of the program's significance to mothers, motivating consistent attendance and minimize dropout rates.
- To investigate how health workers can effectively communicate the value of the intervention (including the provided books and toys) to the mothers, the families and the wider community.

**Guided Questions:**

1. You have a lot of experience raising or caring for young children, so we are interested in your thoughts on early childhood development (ECD) for children under 3 years old.
   - What things are important for promoting young children's development (under 3)?
   - What works best to support their development in the early years?
2. You know the (Buddhi) program implemented through the community clinics. Mothers of children aged between 6-36 months attend fortnightly group sessions at the clinic and the health workers provide mothers with books and toys and demonstrate activities the mothers can do with their children. We know these developmentally focused toys and books offer benefits for young children and can support children's learning and growth.
   - How can we best showcase these benefits to the community?
3. We understand that mothers in the community must juggle various responsibilities like family commitments, chores, and some mothers live further from the clinic. These issues sometimes make it difficult for mothers to attend sessions regularly.
   - How can families work together to overcome these challenges and ensure that mothers can fully participate in this program.
   - What strategies or solutions can you suggest that could encourage and support mothers' consistent attendance?
4. We understand that many mothers and other family members in the community might not be aware of this program and its benefits.
   - What are some challenges you see in raising awareness about the program within the community?
   - What kind of information or support do you think would be most helpful for mothers and other family members to feel motivated to participate in the program regularly?
   - In your opinion, how can health workers more effectively communicate the benefits of the resources and the ways they can be used to enhance children's learning and development?

5. We are eager to improve community awareness about the program. What strategies could we use to encourage mothers and other family members to prioritize attendance and see the long-term value of the program for their children?
   - Beyond traditional methods like posters or flyers, what can be done to encourage mothers and other family members to learn more about the program?

- How can we make sure the message reaches everyone in the community?

6. This program aims to benefit the entire community, and we believe community involvement is crucial for its success.
   - What are some ways you think the community can actively participate and support the Community Clinic in enhancing this program?
   - Who are some other community members, who you think has the influence and credibility to encourage families to participate in this program?

**7.** Imagine you're explaining this program to someone who has never heard of it before. What would you highlight as the most important things for them to know?

**Closing:**

Thank you all for your time. As we near the end of our discussion, please feel free to share any additional insights on raising community awareness and ensuring mother's attendance for the program's successful implementation.
